# SPARSEMODr: Rapid simulations of spatially explicit and stochastic models infectious diseases, including COVID-19

**DOI:** 10.1101/2021.05.13.21256216

**Authors:** Joseph R Mihaljevic, Seth Borkovec, Saikanth Ratnavale, Toby D Hocking, Kelsey E Banister, Joseph E Eppinger, Crystal Hepp, Eck Doerry

## Abstract

Building realistically complex models of infectious disease transmission that are relevant for informing public health is conceptually challenging and requires knowledge of coding architecture that can implement key modeling conventions. For example, many of the models built to understand COVID-19 dynamics have included stochasticity, transmission dynamics that change throughout the epidemic due to changes in host behavior or public health interventions, and spatial structures that account for important spatio-temporal heterogeneities. Here we introduce an R package, `SPARSEMODr`, that allows users to simulate disease models that are stochastic and spatially explicit, including a model for COVID-19 that was useful in the early phases of the epidemic. SPARSEMOD stands for **SPA**tial **R**esolution-**SE**nsitive **M**odels of **O**utbreak **D**ynamics, and our goal is to demonstrate particular conventions for rapidly simulating the dynamics of more complex, spatial models of infectious disease. In this report, we outline the features and workflows of our software package that allow for user-customized simulations. We believe the example models provided in our package will be useful in educational settings, as the coding conventions are adaptable, and will help new modelers to better understand important assumptions that were built into sophisticated COVID-19 models.

## 1 Introduction

The emergence of the SARS-CoV-2 pandemic has reinforced the strong role that mathematical models of disease spread play in understanding pathogen transmission and in designing effective public health interventions (Metcalf *et al*., 2015; Ferguson *et al*., 2020; Tian *et al*., 2020; Saad-Roy *et al*., 2020; Shea *et al*., 2020). Models of infectious disease transmission vary widely in their structural form and complexity (Keeling & Rohani, 2008; Adiga *et al*., 2020). Classical models take the form of ordinary differential equations describing “compartments” of the host and/or pathogen population (e.g., susceptible versus infectious hosts) (Anderson & May, 1979), but even these models can become quite complex, containing numerous equations that might, for instance, account for heterogeneities in the host population or for the progression of pathogen-induced disease through various host classes (e.g., hospitalized individuals). Early in the COVID-19 pandemic, many compartment-style models were developed to address hypotheses of how rapidly SARS-CoV-2 was spreading and what impacts non-pharmaceutical interventions might have (Pan *et al*., 2020; Anderson *et al*., 2020), if and when the virus might become endemic (Lavine *et al*., 2021), and even whether regional climate patterns were likely to influence transmission patterns (Baker *et al*., 2020). Agent-based models, which track the (probabilistic) fate of individuals, were also developed to make short-term forecasts and long-term projections of COVID-19 for public health preparedness (Ferguson *et al*., 2020; Hinch *et al*., 2021; Kerr *et al*., 2021; Cramer *et al*., 2022). Many models nested compartment-style or agent-based models within a spatial framework to incorporate the impacts of spatial contagion and human mobility on COVID-19 dynamics (Yamana *et al*., 2020; Pei *et al*., 2020; Arino, 2022; Zhang *et al*., 2022; Wardle *et al*., 2022). These examples highlight the types of dynamics and assumptions modelers might include when constructing mathematical and computational models of infectious disease transmission, and these choices have important implications for the resulting dynamics.

During the COVID-19 pandemic, some key dynamics have been built into many models that influence the insights that emerge from modeling studies, including short-term forecasts of disease or longer-term projections of system behavior. Arguably some of the most important of these model dynamics and assumptions have been: stochastic transmission processes, such as super-spreading events or probabilistic system behavior; time-varying transmission dynamics due to public health interventions and changes to human behavior (e.g., stay-at-home orders or mask-wearing); and, spatial heterogeneity in transmission, driven in part by human mobility. We will briefly highlight examples of how these dynamics have been important for modeling the spread of SARS-CoV-2, especially in the early phases of the pandemic.

Adding stochastic dynamics to models, such as demographic stochasticity and environmental stochasticity, allows us to capture key processes that are especially impactful early in epidemic behavior and in small populations (Lambert *et al*., 2018). Demographic stochasticity refers to the probabilistic events that befall host individuals, whereas environmental stochasticity refers to random changes to the values of model parameters (e.g., transmission rate) that are due to sources not explicitly included in the model (i.e., environmental sources). Environmental stochasticity is sometimes used to account for heterogeneity in transmission rate among host individuals, such as the impact of super-spreaders (Lloyd-Smith *et al*., 2005), whereas demographic stochasticity accounts for dynamics such as the probabilistic burn-out of a pathogen. For COVID-19, models that included demographic stochasticity where particularly important for explaining disease patterns in regions with small populations (Engbert *et al*., 2021). Moreover, models that included both demographic and environmental stochasticity helped to disentangle the effects of both sources of stochasticity on observed COVID-19 case counts during different phases of the outbreak (Hwang *et al*., 2022). Importantly, accounting for stochastic model behavior also facilitates a more rigorous quantification of uncertainty in model forecasts (Cramer *et al*., 2022). There are many methods to implement stochastic dynamics within models (Allen, 2017), for instance using Gillespie-style algorithms to iterate the differential equations that describe compartment models (Gillespie, 2001; Ganyani *et al*., 2021), or through agent-based modeling, which inherently deals with probabilistic events that befall individual hosts.

Almost all of the models of COVID-19 referenced thus far have needed to deal with the fact that SARS-CoV-2 transmission dynamics and the dynamics of COVID-19 hospitalization changed dramatically throughout the pandemic, due for instance to non-pharmaceutical public health interventions meant to slow transmission, or due to changes in hospital practice to treat and triage COVID-19 patients. Many COVID-19 models estimated time-varying transmission rates from case counts or hospitalization data, or models inferred changing transmission rates using estimates of the pathogen’s instantaneous reproductive number (Cramer *et al*., 2022; Gostic *et al*., 2020; Pei *et al*., 2020). Therefore models need to be flexible enough to allow the user to specify time-varying parameter values. For compartment-style differential equation models, allowing time-varying parameters computationally requires that the models are iterated forward, re-defining parameter values at appropriate time-points.

Modeling spatially explicit disease dynamics is important for testing hypotheses about the roles of human mobility on spatial contagion, the effectiveness of interventions that impact movement, and understanding the sources of transmission within and among regions. Spatial models also allow for more refined forecasts within specific locations. Indeed, spatial models are often necessary to explain large-scale patterns of disease transmission that cannot be captured by simpler, non-spatial models (Eggo *et al*., 2021). There are several approaches towards spatially explicit modeling (Riley, 2007), including agent-based network models that situate individual hosts in specific spatial positions (Ferguson *et al*., 2020; Kerr *et al*., 2021; Hinch *et al*., 2021), or partial differential equations that assume hosts can move continuously across spatial dimensions. One of the most popular approaches, however, is meta-population disease modeling. In meta-population models of disease, distinct host populations are explicitly situated geographically such that movement of host individuals between the populations affects local and regional transmission dynamics (Rohani *et al*., 1999; Lachiany & Stone, 2012; Ferrari *et al*., 2010). Important basic theory of spatial epidemiology has emerged from the analysis of meta-population models (Arino, 2009; Wang & Li, 2014), which has in turn popularized the use of these spatial models for understanding and forecasting real public health threats (Kraemer *et al*., 2019). Many early analyses of COVID-19 used meta-population models to account for spatial heterogeneities in transmission and disease outcomes (Pei *et al*., 2020; Yamana *et al*., 2020; Gatto *et al*., 2020; Yang *et al*., 2021; Zebrowski *et al*., 2021; Arino, 2022). The most complex meta-population models of COVID-19 parameterize human movement based on an emerging suite of mobility data, often derived from mobile devices (Hou *et al*., 2021; Wardle *et al*., 2022; Hu *et al*., 2021).

Here, we introduce an R package, `SPARSEMODr`, that includes two illustrative examples of spatially-explicit and stochastic meta-population disease models, including one of COVID-19. Some agent-based simulation models of COVID-19 have been introduced that develop software for users to experiment with the models (Kerr *et al*., 2021; Hinch *et al*., 2021), and several published models provide openly available code for users to download (Cramer *et al*., 2022; Ferguson *et al*., 2020). Our package focuses on meta-population models in which users can simulate spatial and temporal heterogeneities in transmission rates, effects of host movement, and other user-defined dynamics that influence local and regional patterns of disease. The COVID-19 model that we provide was used by our group and others (Gel *et al*., 2020) to characterize spatial and temporal variation in transmission patterns and disease outcomes (e.g., hospitalizations and ICU admissions) in Arizona, USA. While the model is no longer complex enough to capture many of the current critical aspects of COVID-19 transmission and disease (e.g., the model does not include age-structure, vaccination, nor the circulation of multiple variants), the models in `SPARSEMODr` should nonetheless be useful for educational purposes. Importantly, the package demonstrates some specific conventions of coding stochastic meta-population models that could easily be carried over to different host-pathogen systems in teaching, research, or applied contexts.

## 2 Available models

Currently we offer two models in the `SPARSEMODr` package: one is a more classic Susceptible-Exposed-Infectious-Removed (SEIR) model, and the other is an SEIR-style model that more specifically describes the transmission of SARS-CoV-2 and COVID-19 progression from exposure through hospitalization through mortality. Both models are compartmental disease models that are simulated within a meta-population context, which we describe below. We supply detailed vignettes that describe different use-cases of the `SPARSEMODr` package, which can be found on CRAN: https://cran.r-project.org/web/packages/SPARSEMODr/index.html and the package GitHub repository.

The SEIR model is described by the following set of ordinary differential equations:

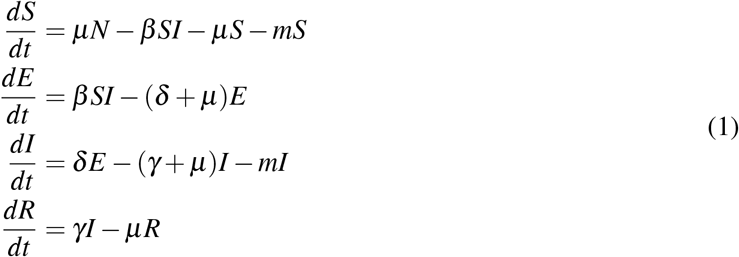

Here, the state variables represent the numbers of hosts in each class (e.g., *S* is the number of susceptible hosts). Following some classic conventions, the model assumes that all host classes can reproduce at rate *µ*, which is equal to the natural death rate. This has the effect of maintaining total host population size static through time. Moreover, all offspring enter the susceptible class, such that *N* = *S* + *E* + *I* + *R*. The pathogen incubates within exposed individuals for an average latency time of 1*/δ*, and infectious individuals recover at rate *γ*. This model structure leads to exponentially distributed sojourn times. As we will describe below, we assume that susceptible and infectious hosts can commute away from their focal population at emigration rate, *m*, but hosts return after one day. This commuter-style movement allows susceptible individuals to become exposed by infectious individuals in other populations and infectious individuals to spread the pathogen to other populations. An example simulation of the model, in which we impose sinusoidal forcing of the transmission rate, is shown in Figure 1.

**Figure 1:**
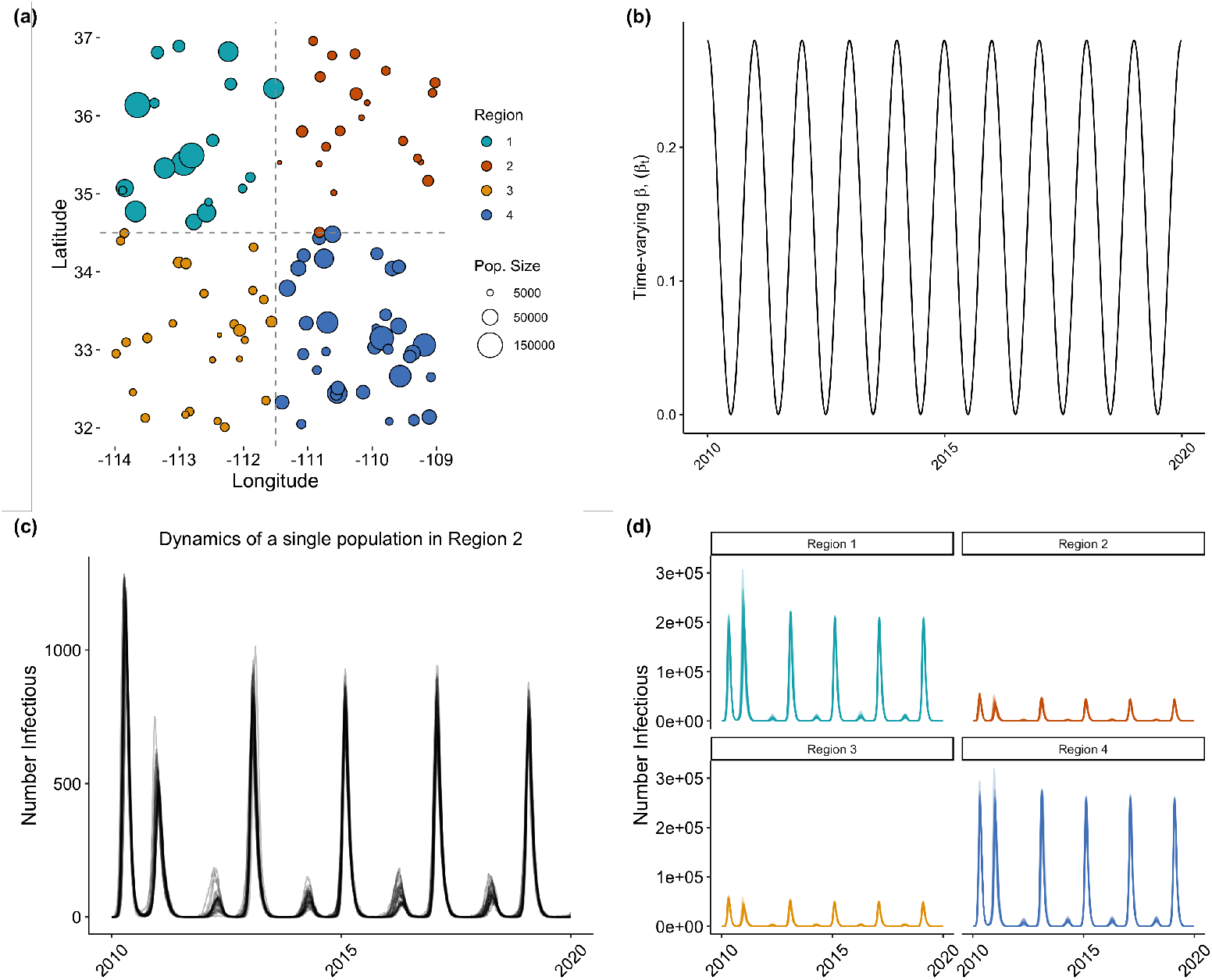
Simulation of the `SPARSEMODr` SEIR model. (a) Simulated host meta-population. (b) Imposing a sinusoidal pattern of time-varying transmission rate, *β*_*t*_. (c) Pattern of number infectious over time from a single sub-population. The lighter lines are individual realizations of the stochastic model. (d) Aggregating local patterns to regional scales to explore differences in emergent patterns.

The COVID-19 model is described by the following equations (Fig. 2; see also Gel *et al*. (2020)):

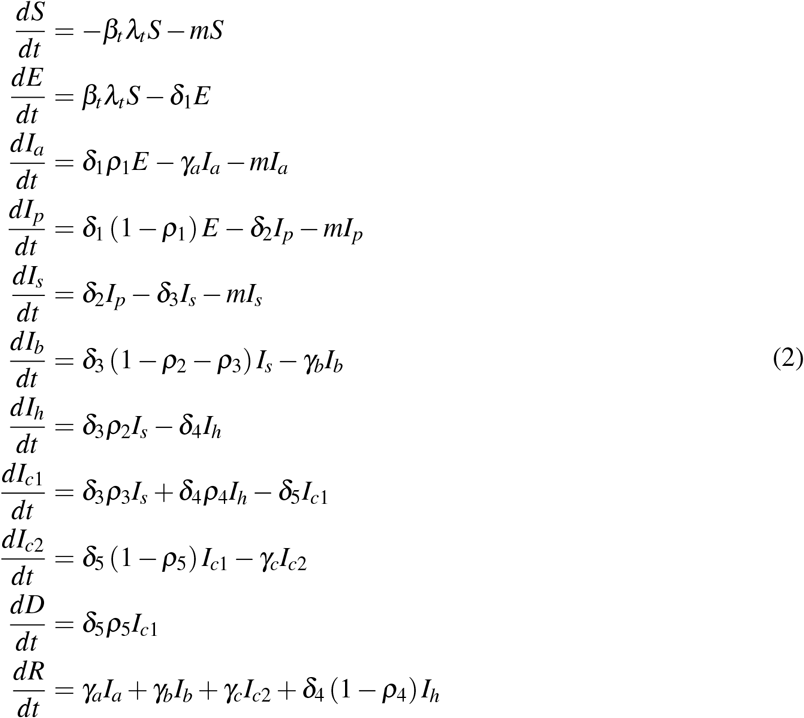

**Figure 2:**
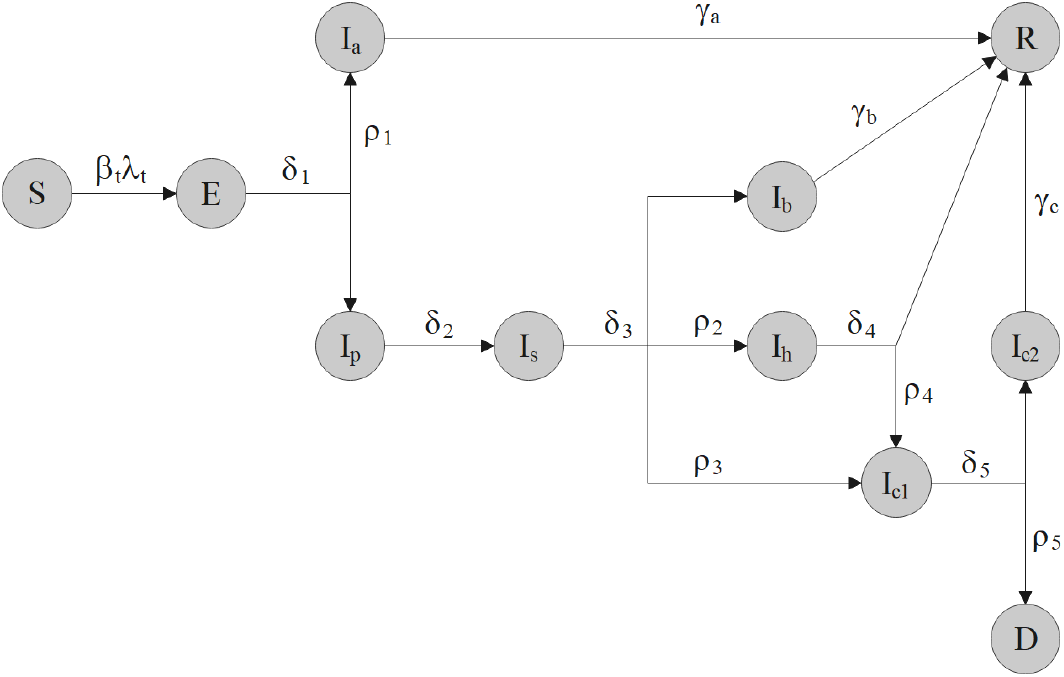
COVID-19 model schematic. State variables and parameters are defined in Tables 1 and 2.

The term *λ*_*t*_ represents a component of the force of infection, given by:

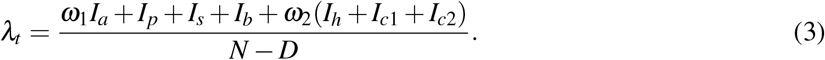

**Table 1:** COVID-19 model state variables

| State Variable | Description | Corresponding model input |
| --- | --- | --- |
| $S$ | Number of susceptible individuals | input_S_pops |
| $E$ | Number of exposed individuals | input_E_pops |
| $I_a$ | Number of asymptomatic individuals | input_I_asym_pops |
| $I_p$ | Number of pre-symptomatic individuals | input_I_presym_pops |
| $I_s$ | Number of mildly symptomatic individuals | input_I_sym_pops |
| $I_b$ | Number of mildly symptomatic individuals on bed rest at home | input_I_home_pops |
| $I_h$ | Number of hospitalized individuals | input_I_hosp_pops |
| $I_{c1}$ | Number of individuals in the ICU | input_I_icu1_pops |
| $I_{c2}$ | Number of individuals in the recovery (step-down) ICU | input_I_icu2_pops |
| $D$ | Number of deceased individuals | input_D_pops |
| $R$ | Number of recovered individuals | input_R_pops |

**Table 2:**
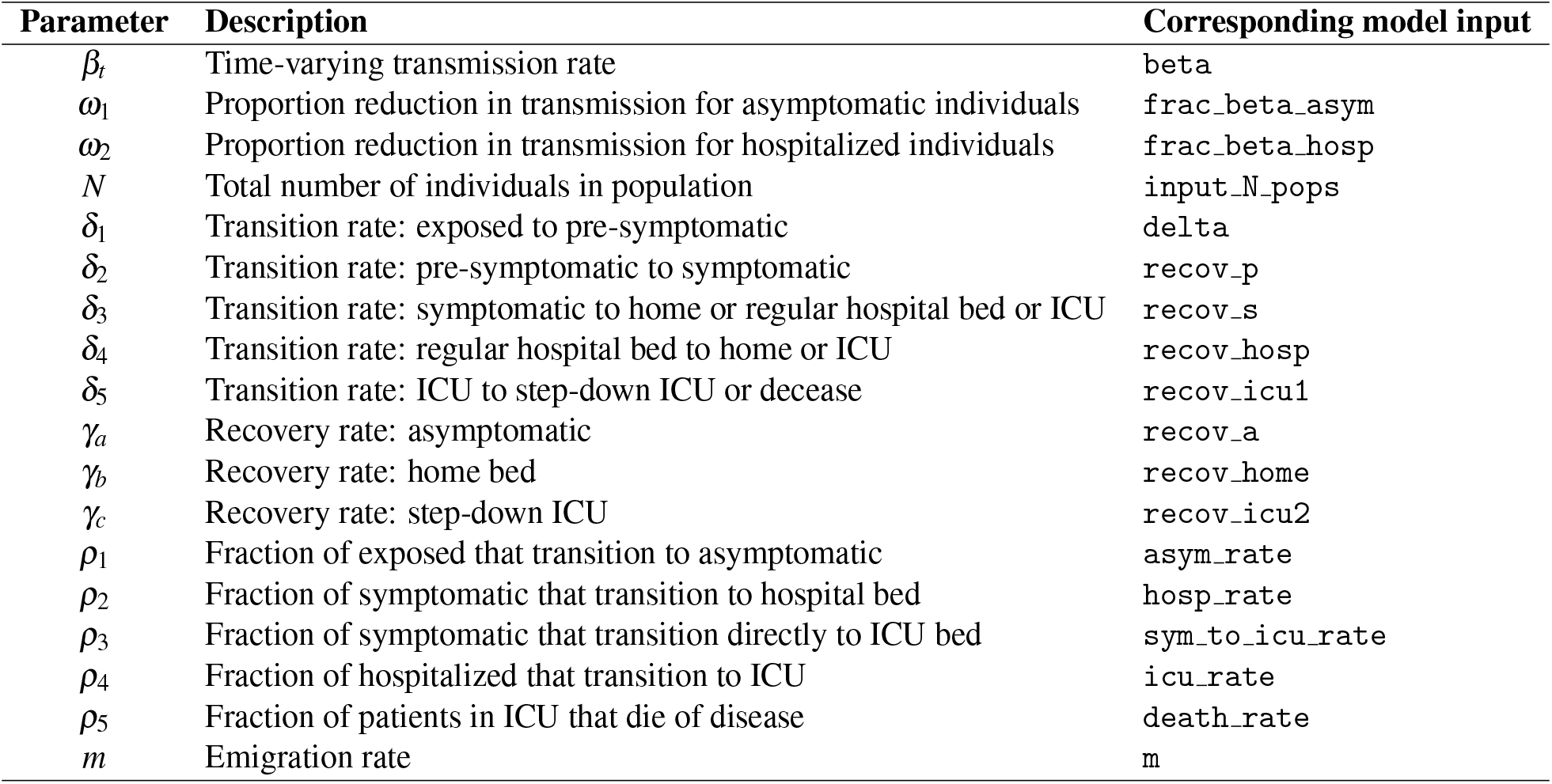
COVID-19 model parameter descriptions

Definitions of state variables and model parameters are shown in Tables 1 and 2. Note that *N* represents the total number of hosts in the population. We assume that individuals in the *S, I*_*a*_, *I*_*p*_, and *I*_*s*_ compartments can move between populations at emigration rate, *m*. We focus on these host classes as they can influence local and regional transmission dynamics, whereas, for example, commuter-style movement of exposed individuals would have no effect.

In the package manual, we provide default parameter values, but all of the COVID-19 model parameters can be user-specified. The package also conducts parameter validation steps via warnings and errors to ensure specified parameters are within feasible limits. Figure 3 shows a simulated example using the COVID-19 model.

**Figure 3:**
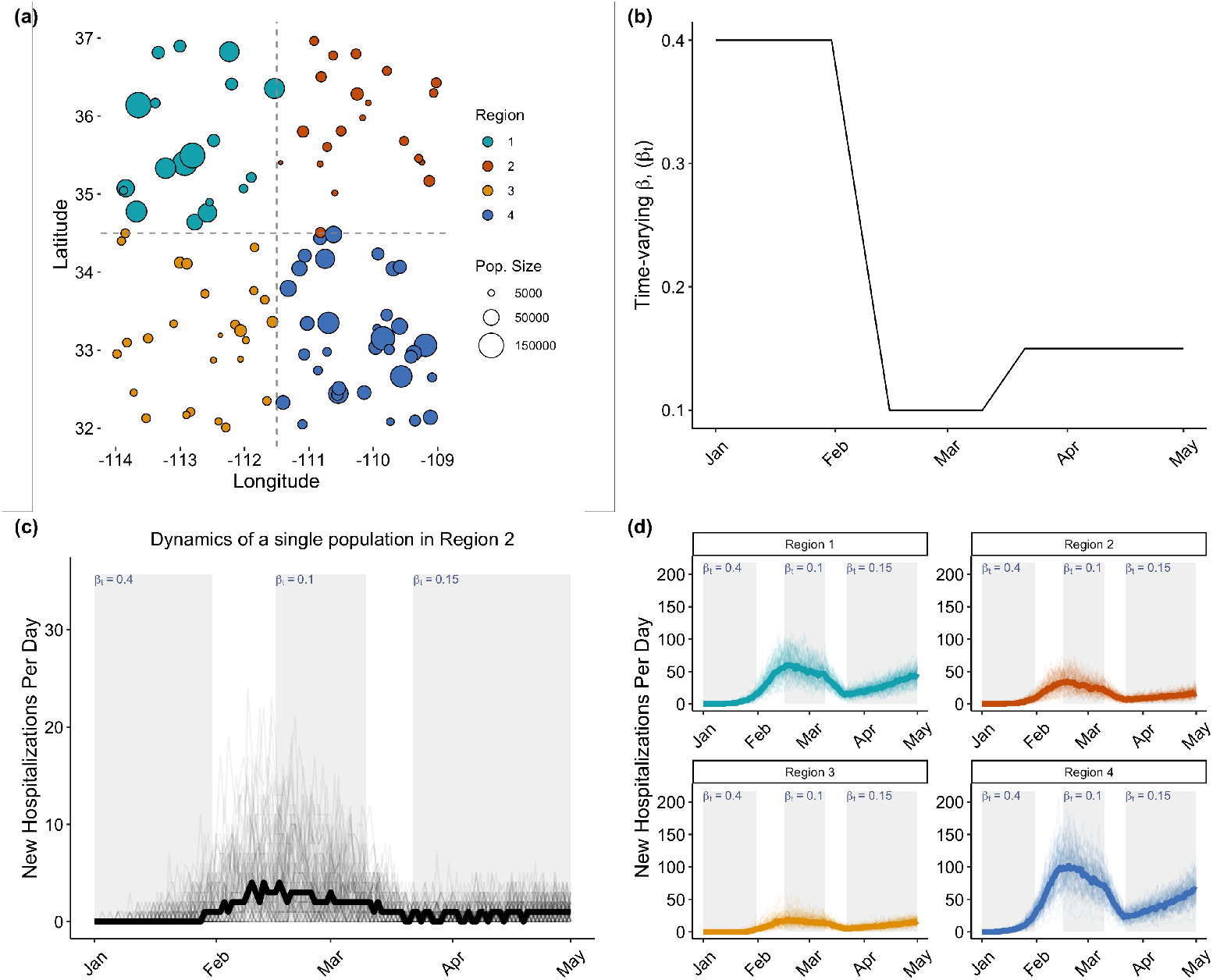
Simulation of the `SPARSEMODr` COVID-19 model. (a) Simulated host meta-population. (b) Imposing a pattern of time-varying transmission rate, *β*_*t*_. (c) Pattern of new hospitalizations over time from a single sub-population. The lighter lines are individual realizations of the stochastic model, while the dark line is the median trajectory of these realizations. (d) Aggregating local patterns to regional scales to explore differences in emergent patterns. Light and dark lines as in panel (c).

## 3 Key features of SPARSEMODr disease models

We implement a common coding architecture for the two models provided in `SPARSEMODr` to highlight particular modeling conventions that are useful to consider in theoretical and applied modeling studies. Here we describe these key features of the spatially-explicit and stochastic disease models. The models in `SPARSEMODr` are coded in C++ and use the `Rcpp` package to conveniently wrap the functions into the R computing environment (Eddelbuettel, 2013), integrating the speed of C++ and the user-friendliness of R. The output of each model is a data frame that includes the value of each state variable and the number of new “events” (e.g., new exposures, new hospitalizations) per time step per population per stochastic realization of the model.

### 3.1 Stochastic dynamics

#### Demographic stochasticity

The models include demographic stochasticity, which implements the effects of probabilistic events that befall individuals in a population and that can affect epidemic trajectories. We use an event-driven approach in which the differential equations are iterated forward in daily time steps using a Gillespie-style algorithm known as the tau-leaping algorithm (Gillespie, 2001). The tau-leaping algorithm is more flexible and more computationally efficient compared to several other simulation and numerical integration techniques (Ganyani *et al*., 2021). And, because this form of demographic stochasticity requires us to use integers for the host classes (i.e., numbers of hosts in each class), it allows users to track the number of new events occurring per time step.

#### Stochastic transmission process

We also implement daily stochastic variation in the transmission rate, a form of environmental stochasticity. We assume that as the number of infectious individuals increases, the variation in transmission rate decreases, emphasizing that stochasticity has larger effects in smaller populations (Keeling & Rohani, 2008). Our specific form of stochasticity implies that there is heterogeneity in the transmission rate among individuals in the host population, such that our methods can account for super-spreader events, for instance, which have disproportionate effects early in the epidemic. The functional form of stochasticity is:

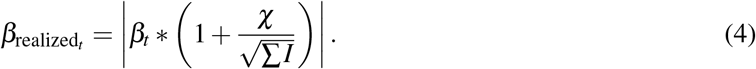

For every time step *t* we draw a random variate, *χ*, from a normal distribution with a mean of zero and a standard deviation of *σ*, such that *χ ~ N*(0, *σ*), and *σ* is a user-controlled model input, `stoch_sd`. We calculate the total number of infectious individuals in the population, which we abbreviate ∑ *I*, because the summation will depend on the model and which state variables represent infectious individuals (e.g., there are several infectious states in the COVID-19 model). The vertical bars show that 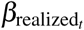 is the absolute value of the right-hand expression.

### 3.2 Spatial, meta-population dynamics

To iterate the `SPARSEMODr` models in a spatial context, we embed the equations into sub-populations that make up a meta-population, in which migration connects sub-populations. Migration between populations in the meta-population thus affects local and regional transmission dynamics. Susceptible individuals in a focal population can become exposed to the pathogen by infectious “visitors” from other sub-populations or by infectious visitors from outside of the meta-population (“immigrants”). Similarly, susceptible individuals can visit a different sub-population and become exposed by the resident infectious individuals. We model movement using a commuter-style approach, in which hosts can move to an alternative sub-population during a time step, but then they return to their home sub-population before the start of the next time step. After we move individuals to new populations, transmission between susceptible residents and infectious visitors is determined, and the visitors then leave the population. In other words, in a single day (i.e., time-step), transmission first occurs within a population and then additional transmission can occur after hosts commute.

The user controls the per-capita emigration rate, *m*, of susceptible and infectious hosts. Our models assume that the probability of moving to any specific population is dictated by a simple, distance-based dispersal kernel:

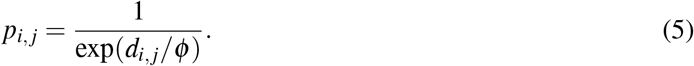

Here *p*_*i, j*_ is the probability of moving from population *j* to population *i*, and *d*_*i, j*_ is the euclidean distance between the two populations. Larger values of the *ϕ* parameter (controlled as user input `dist_phi`) makes it more likely for hosts to travel farther distances. To determine the population to which migrants will move during any given time step, we draw from a multinomial probability distribution using the *p*_*i, j*_ values.

The models also allow the effects of immigrants, who do not usually reside in the meta-population, to commute to the meta-population each day (e.g., out-of-state tourists). The user can define the model input `imm_frac`, which is the proportion of the focal population that may constitute visitors on any given day. For example, if for a given focal population the population size is 1000 hosts, and `imm_frac` = 0.05, an average of 50 immigrants may arrive on a given day. The exact number of these immigrants arriving on a given day is determined by drawing from a Poisson distribution. Then, the number of infectious visitors from this pool of immigrants is assumed to be proportional to the number of infectious residents in the focal population. In other words, we assume that the pathogen is present in “non-resident” populations at similar prevalence as the focal population.

### 3.3 Time-varying parameters

The `SPARSEMODr` models allow users to specify certain time-varying parameters, which take on unique values per day. Both models allow the transmission rate, *β*_(*t*)_, to vary over time, and we allow the user to control how movement dynamics change over time with time-varying values of *m, ϕ*, and the fraction of the population that constitutes outside immigrants, `imm_frac`. In the COVID-19 model, we also allow time-varying fraction hospitalized, *ρ*_2_, transition rate of hospitalized individuals, *δ*_4_, fraction advancing to the ICU, *ρ*_4_, and fraction of ICU patients that die of disease, *ρ*_5_. We chose to allow these parameters to vary over time to emphasize that these rates, in reality, very likely changed throughout the COVID-19 pandemic, for instance, due to changes in hospital policies and improvements to patient care. The time-varying parameters can be specified in one of two ways, which we demonstrate in the **Work flow** section below.

### 3.4 Frequency- and density-dependent transmission

In the `SPARSEMODr` models, the transmission process can be described as frequency-dependent, where contact rates are invariable to population density, or density-dependent, where contact rates depend on population density (Hopkins *et al*., 2020). For frequency-dependent transmission, we divide the user-specified value of transmission rate by the total number of hosts within a given sub-population. For example, in the SEIR model, we have the following expression describing mass-action transmission per sub-population, 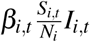, where *β*_*i,t*_ is the user-specified transmission rate for sub-population *i* at time step *t*, and *N*_*i*_ is the total host population size for sub-population *i*. Therefore, with frequency-dependent transmission, the effect of a single infectious host on the risk of infection is modulated by the fraction of the host population that is still susceptible, irrespective of the host population density (i.e., number of hosts per unit area) within that sub-population.

Alternatively, for density-dependent transmission, the user can specify a non-linear Monod equation that describes the relationship between host population density and the transmission rate. The Monod equation is:

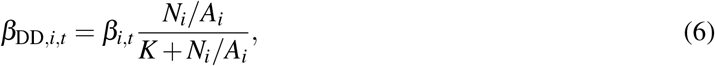

Here, *β*_*i,t*_ is the transmission rate supplied by the user, but importantly this functionally becomes the maximum possible transmission rate for sub-population *i* at time step *t*. Thus, *β*_DD,*i,t*_ is the density-dependent transmission rate for sub-population *i* at time step *t. N*_*i*_*/A*_*i*_ is the density of the focal host population (i.e., number of hosts, *N*_*i*_ per unit area, *A*_*i*_). To use this form of transmission, the area per population, *A*_*i*_, must therefore be specified by the user as input vector, `census_area`. Finally, *K* is a constant that controls the effect of density on the transmission rate. More specifically, *K* is the half-saturation constant at which point *β*_DD,*i,t*_*/β*_*i,t*_ = 0.5. In general, when *K* = 0, there is no effect of density on transmission rate, but larger values of *K* mean that low host densities more strongly reduce the transmission rate (i.e., transmission rate more strongly depends on host population density). The value of *K* is user-controlled by model input, `dd_trans_monod_k`.

## 4 Work flow

### 4.1 Process overview

`SPARSEMODr` runs spatial, stochastic models across a user-specified grid of populations. In Figures 1 and 3, we simulated populations that are scattered across a spatial lattice; one could imagine these are local communities situated within counties, labelled as “Regions”. In these particular simulations, we imposed a time-varying transmission rate, *β*_*t*_, but we assumed that each local population experiences the same pattern of time-varying transmission. The models track the spread of the disease in each sub-population, such that we can computationally aggregate patterns from local populations to higher spatial scales (e.g., “Regions”) to look at emergent patterns. Below we describe the necessary setup and declarations to run these simulations for the models in `SPARSEMODr`, focusing on the more complex COVID-19 model.

### 4.2 Declaring initial conditions and constant parameters

Users must specify the initial conditions of the state variables. To initiate the COVID-19 model, at least one of the following vectors must be supplied with a value greater than one for at least one sub-population:

`input_E_pops`, `input_I_asym_pops`, `input_I_presym_pops`, `input_I_sym_pops`, `input_I_home_pops`, `input_I_hosp_pops`, `input_I_icu1_pops`, or `input_I_icu2_pops`. If any initial values of state variables are not supplied, they are assumed to be zero. Moreover, either a vector of `input_N_pops` or a vector of `input_S_pops` must be supplied. As an example, we will specify:

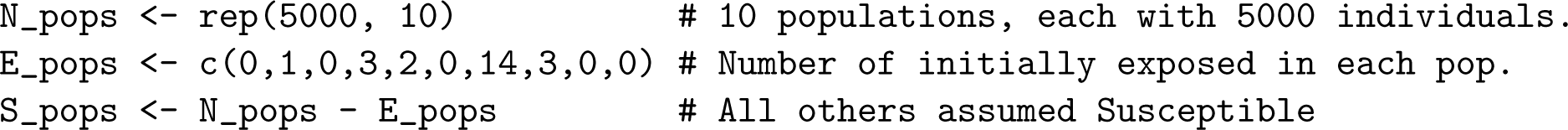

The user must also declare some inputs that are shared between the `SPARSEMODr` models: `input_dist_mat` is a matrix that specifies the distance between populations; `input_realz_seeds` is a vector of integer values to seed the random realizations of the model; `stoch_sd` is the standard deviation of the stochastic transmission rate, *σ*, as described above; `trans_type` specifies the type of transmission type (frequency- or density-dependent); `input census_area` is the spatial area of each population, but this is only required when transmission is declared as density-dependent; `dd_trans_monod_k` the parameter controlling density’s effect on transmission, again only required for density-dependent transmission; and `input_tw` a time window object, which we will describe in the next section.

The next step is to populate the model-specific “control” object, which is a special R class defined in our package. Each model has its own control class that includes the model-specific parameters and initial conditions of the state variables.

As a simple example with the COVID-19 model, given the initial conditions above, here we will use default parameter values for all but two parameters:

~~~
my_covid19_control <- SPARSEMODr::covid19_control(
 input_S_pops = S_pops, # susceptible population counts
 input_E_pops = E_pops, # exposed population counts
 asym_rate = 0.4,       # fraction of exposed that become asymptomatic
 recov_icu1 = 0.125)    # average ICU recovery rate, i.e., 8 days (1/8)
~~~

Also, because `input_N_pops` was not provided, the package internally assumes `input_N_pops` = `input_S_pops` + `input_E_pops`. `covid19_control()` returns a named list of vectors that must be supplied when running the model.

### 4.3 Declaring time-varying parameters with the time-windows object

A `time windows` object is required to specify the time-varying parameters (or whether these parameters are assumed constant). There are two ways to specify time windows: (1) specifying values for each time step in the simulation (i.e., “daily” method), or (2) the starting and ending dates of user-defined time windows. For the “daily” method, the parameter vectors must be of size equal to the number of days in the simulation. For the start date/end date method, values for each parameter are assigned at the beginning and end of the time window. Then, the back-end code calculates a linear interpolation to assign daily values of the parameter; in other words, the parameter values change linearly from the starting value to the ending value over the number of days within the time window.

In the following example, we specify the start and end dates of our time windows for time-varying transmission rate, to match the pattern shown in Fig. 3(b). Note that we format dates using the `lubridate` package (Grolemund & Wickham, 2011), although other methods are acceptable as long as they are objects of class `Date` in R.

~~~
# Function to specify all of the required parameters in a single time-window
# In this example, only r0 is variable between the time periods
one.window <- function(
 start_dates,   #start of time window (date class)
 end_dates,     #end of time window (date class)
 beta,          #value of time-varying beta
 dist_phi=150,  #controls shape of dispersal kernel
 m=0.1,         #per-capita movement rate, i.e., move every 10 days on avg.
 imm_frac=0){   #zero outside immigration
 data.frame(beta, start_dates, end_dates, dist_phi, m, imm_frac)
}
library(lubridate) # for mdy()
time.window.args <- rbind(# Specify the components of 5 time windows
 one.window(mdy(“1-1-20”), mdy(“1-31-20”), beta=0.40),
 one.window(mdy(“2-1-20”), mdy(“2-15-20”), beta=0.10),
 one.window(mdy(“2-16-20”), mdy(“3-10-20”), beta=0.10),
 one.window(mdy(“3-11-20”), mdy(“3-21-20”), beta=0.15),
 one.window(mdy(“3-22-20”), mdy(“5-1-20”), beta=0.15)
)
# Populate the required object of class time_windows
my_tw <- do.call(SPARSEMODr::time_windows, time.window.args)
~~~

The package documentation and the vignettes give additional examples of how to flexibly structure these time window objects. Notably, the vignettes demonstrate how to specify time-varying *β*_*t*_ values for each sub-population individually, to explore effects of spatially heterogeneous transmission rates within the meta-population. In this latter case, users would specify a list of `beta` vectors.

### 4.4 Running stochastic realizations in parallel

For efficiency and to reduce run time, we suggest simulating stochastic realizations of the `SPARSEMODr` models in parallel. We have therefore created a function, `model_parallel()`, that leverages the `future` package (Bengtsson, 2020) and compiles the output of the individual model runs into a user-friendly data frame. As an example:

~~~
# Specify the number of realizations to run:
n_realz <- 75
# Specify unique seeds to run the realizations.
## Note, realizations with the same seeds will produce
## equivalent output
my_realz_seeds <- 1:n_realz
# Run the model in parallel and store the output:
model_output <- SPARSEMODr::model_parallel(
                          input_dist_mat = dist_mat,
                          input_census_area = census_area, input_tw = my_tw,
                          input_realz_seeds = my_realz_seeds, control = my_covid19_control,
                          …universal_model_params…)
~~~

Here “… `universal_model_params` … “ represents the universal model parameters, such as `trans_type` or `stoch_sd`, as described above. Note that `dist_mat` is a pairwise distance matrix, and `census_area` is a vector of population areas (e.g., in km^2^). The package documentation and vignettes show examples of creating these two inputs. Since `model_parallel()` produces a data frame, the output can easily be subset and summarized for plotting.

## 5 Conclusion

The `SPARSEMODr` package illustrates coding paradigms to specify and simulate complex disease models that are both stochastic and spatially explicit. In pedagogical contexts, `SPARSEMODr` models can be used to demonstrate the effects of time-varying transmission rates (e.g., as affected by disease interventions), demographic and environmental stochasticity, frequency-versus density-dependent transmission, and spatial heterogeneities. For instance, simulating the `SPARSEMODr` SEIR model can reinforce foundational theoretical concepts in mathematical epidemiology, such as spreading waves of disease or spatial (a)synchrony of epidemics. The `SPARSEMODr` COVID-19 model can be used to help students understand the key features of models built early in the pandemic to address public health issues, for example, highlighting the effects of public health interventions, spatiotemporal effects of altering human movement, or the changing landscape of hospitalization dynamics before vaccines were available. Because we develop a standard coding architecture for the two models in `SPARSEMODr`, we can envision multiple future package developments. For instance, our team or outside contributors can add models that follow the same coding standards, such as vector-borne disease models, or COVID-19 models that include additional layers of realism, such as age-structure, vaccination, and the dynamics of co-circulating variants. For package contributors, a new model would have to follow the current conventions, which can be copied from our GitHub repository (https://github.com/NAU-CCL/SPARSEMODr). Our team would approve all outside contributed model structures by forking and merge requests. We also plan to modularize the `SPARSEMODr` coding conventions using APIs so that they can be auto-populated for new models, reducing the need for copy-and-pasting code. We hope that the community of disease modelers and educators will help us to refine the package and to make it more accessible to broader audiences.

## Data Availability

No data used

## 7 Author Contributions

JRM, CH, and ED designed the project. JRM, TDH, SB, SR, KEB, and JEE contributed code and methods. JRM, TDH, and SB designed the R package structure. JRM, SB, and SR wrote the first draft of this manuscript, and all authors contributed to manuscript revisions.

## 8 Acknowledgements

This material is based upon work supported by the National Science Foundation under Grant No. 2028629.

## References

Adiga, A., Dubhashi, D., Lewis, B., Marathe, M., Venkatramanan, S. & Vullikanti, A. (2020) Mathematical models for covid-19 pandemic: A comparative analysis. Journal of the Indian Institute of Science, 100, 793–807.

Allen, L.J. (2017) A primer on stochastic epidemic models: Formulation, numerical simulation, and analysis. Infectious Disease Modelling, 2, 128–142.

Anderson, R.M. & May, R.M. (1979) Population biology of infectious diseases: Part i. Nature, 280, 361–367.

Anderson, R.M., Heesterbeek, H., Klinkenberg, D. & Hollingsworth, T.D. (2020) How will country-based mitigation measures influence the course of the covid-19 epidemic? The Lancet, 395, 931–934.

Arino, J. (2009) Diseases in metapopulations. Modeling and Dynamics of Infectious Diseases, pp. 64–122.

Arino, J. (2022) Describing, modelling and forecasting the spatial and temporal spread of covid-19: A short review. Mathematics of Public Health, pp. 25–51.

Baker, R.E., Yang, W., Vecchi, G.A., Metcalf, C.J.E. & Grenfell, B.T. (2020) Susceptible supply limits the role of climate in the early sars-cov-2 pandemic. Science, p. eabc2535.

Bengtsson, H. (2020) future: Unified Parallel and Distributed Processing in R for Everyone. R package version 1.18.0.

Cramer, E.Y., Ray, E.L., Lopez, V.K., Bracher, J., Brennen, A., Rivadeneira, A.J.C., Gerding, A., Gneiting, T., House, K.H., Huang, Y., Jayawardena, D., Kanji, A.H., Khandelwal, A., Le, K., Mühlemann, A., Niemi, J., Shah, A., Stark, A., Wang, Y., Wattanachit, N., Zorn, M.W., Gu, Y., Jain, S., Bannur, N., Deva, A., Kulkarni, M., Merugu, S., Raval, A., Shingi, S., Tiwari, A., White, J., Abernethy, N.F., Woody, S., Dahan, M., Fox, S., Gaither, K., Lachmann, M., Meyers, L.A., Scott, J.G., Tec, M., Srivastava, A., George, G.E., Cegan, J.C., Dettwiller, I.D., England, W.P., Farthing, M.W., Hunter, R.H., Lafferty, B., Linkov, I., Mayo, M.L., Parno, M.D., Rowland, M.A., Trump, B.D., Zhang-James, Y., Chen, S., Faraone, S.V., Hess, J., Morley, C.P., Salekin, A., Wang, D., Corsetti, S.M., Baer, T.M., Eisenberg, M.C., Falb, K., Huang, Y., Martin, E.T., McCauley, E., Myers, R.L., Schwarz, T., Sheldon, D., Gibson, G.C., Yu, R., Gao, L., Ma, Y., Wu, D., Yan, X., Jin, X., Wang, Y.X., Chen, Y., Guo, L., Zhao, Y., Gu, Q., Chen, J., Wang, L., Xu, P., Zhang, W., Zou, D., Biegel, H., Lega, J., McConnell, S., Nagraj, V.P., Guertin, S.L., Hulme-Lowe, C., Turner, S.D., Shi, Y., Ban, X., Walraven, R., Hong, Q.J., Kong, S., van de Walle, A., Turtle, J.A., Ben-Nun, M., Riley, S., Riley, P., Koyluoglu, U., DesRoches, D., Forli, P., Hamory, B., Kyriakides, C., Leis, H., Milliken, J., Moloney, M., Morgan, J., Nirgudkar, N., Ozcan, G., Piwonka, N., Ravi, M., Schrader, C., Shakhnovich, E., Siegel, D., Spatz, R., Stiefeling, C., Wilkinson, B., Wong, A., Cavany, S., España, G., Moore, S., Oidtman, R., Perkins, A., Kraus, D., Kraus, A., Gao, Z., Bian, J., Cao, W., Ferres, J.L., Li, C., Liu, T.Y., Xie, X., Zhang, S., Zheng, S., Vespignani, A., Chinazzi, M., Davis, J.T., Mu, K., y Piontti, A.P., Xiong, X., Zheng, A., Baek, J., Farias, V., Georgescu, A., Levi, R., Sinha, D., Wilde, J., Perakis, G., Bennouna, M.A., Nze-Ndong, D., Singhvi, D., Spantidakis, I., Thayaparan, L., Tsiourvas, A., Sarker, A., Jadbabaie, A., Shah, D., Penna, N.D., Celi, L.A., Sundar, S., Wolfinger, R., Osthus, D., Castro, L., Fairchild, G., Michaud, I., Karlen, D., Kinsey, M., Mullany, L.C., Rainwater-Lovett, K., Shin, L., Tallaksen, K., Wilson, S., Lee, E.C., Dent, J., Grantz, K.H., Hill, A.L., Kaminsky, J., Kaminsky, K., Keegan, L.T., Lauer, S.A., Lemaitre, J.C., Lessler, J., Meredith, H.R., Perez-Saez, J., Shah, S., Smith, C.P., Truelove, S.A., Wills, J., Marshall, M., Gardner, L., Nixon, K., Burant, J.C., Wang, L., Gao, L., Gu, Z., Kim, M., Li, X., Wang, G., Wang, Y., Yu, S., Reiner, R.C., Barber, R., Gakidou, E., Hay, S.I., Lim, S., Murray, C., Pigott, D., Gurung, H.L., Baccam, P., Stage, S.A., Suchoski, B.T., Prakash, B.A., Adhikari, B., Cui, J., Rodríguez, A., Tabassum, A., Xie, J., Keskinocak, P., Asplund, J., Baxter, A., Oruc, B.E., Serban, N., Arik, S.O., Dusenberry, M., Epshteyn, A., Kanal, E., Le, L.T., Li, C.L., Pfister, T., Sava, D., Sinha, R., Tsai, T., Yoder, N., Yoon, J., Zhang, L., Abbott, S., Bosse, N.I., Funk, S., Hellewell, J., Meakin, S.R., Sherratt, K., Zhou, M., Kalantari, R., Yamana, T.K., Pei, S., Shaman, J., Li, M.L., Bertsimas, D., Lami, O.S., Soni, S., Bouardi, H.T., Ayer, T., Adee, M., Chhatwal, J., Dalgic, O.O., Ladd, M.A., Linas, B.P., Mueller, P., Xiao, J., Wang, Y., Wang, Q., Xie, S., Zeng, D., Green, A., Bien, J., Brooks, L., Hu, A.J., Jahja, M., McDonald, D., Narasimhan, B., Politsch, C., Rajanala, S., Rumack, A., Simon, N., Tibshirani, R.J., Tibshirani, R., Ventura, V., Wasserman, L., O’Dea, E.B., Drake, J.M., Pagano, R., Tran, Q.T., Ho, L.S.T., Huynh, H., Walker, J.W., Slayton, R.B., Johansson, M.A., Biggerstaff, M. & Reich, N.G. (2022) Evaluation of individual and ensemble probabilistic forecasts of covid-19 mortality in the united states. Proceedings of the National Academy of Sciences, 119.

Eddelbuettel, D. (2013) Seamless R and C++ Integration with Rcpp. Springer, New York. ISBN 978-1-4614-6867-7.

Eggo, R.M., Dawa, J., Kucharski, A.J. & Cucunuba, Z.M. (2021) The importance of local context in covid-19 models. Nature Computational Science, 1, 6–8.

Engbert, R., Rabe, M.M., Kliegl, R. & Reich, S. (2021) Sequential data assimilation of the stochastic seir epidemic model for regional covid-19 dynamics. Bulletin of Mathematical Biology, 83.

Ferguson, N., Laydon, D., Nedjati Gilani, G., Imai, N., Ainslie, K., Baguelin, M., Bhatia, S., Boonyasiri, A., Cucunuba Perez, Z., Cuomo-Dannenburg, G. et al. (2020) Report 9: Impact of non-pharmaceutical interventions (npis) to reduce covid19 mortality and healthcare demand.

Ferrari, M.J., Djibo, A., Grais, R.F., Bharti, N., Grenfell, B.T. & Bjornstad, O.N. (2010) Rural-urban gradient in seasonal forcing of measles transmission in Niger. Proceedings of the Royal Society B: Biological Sciences, 277, 2775–2782.

Ganyani, T., Faes, C. & Hens, N. (2021) Simulation and analysis methods for stochastic compartmental epidemic models. Annual Review of Statistics and Its Application, 8, 69–88.

Gatto, M., Bertuzzo, E., Mari, L., Miccoli, S., Carraro, L., Casagrandi, R. & Rinaldo, A. (2020) Spread and dynamics of the covid-19 epidemic in italy: Effects of emergency containment measures. 117.

Gel, E.S., Jehn, M., Lant, T., Muldoon, A.R., Nelson, T. & Ross, H.M. (2020) Covid-19 healthcare demand projections: Arizona. PloS one, 15, e0242588.

Gillespie, D.T. (2001) Approximate accelerated stochastic simulation of chemically reacting systems. The Journal of Chemical Physics, 115, 1716–1733.

Gostic, K.M., Mcgough, L., Baskerville, E., Abbott, S., Joshi, K., Tedijanto, C., Kahn, R., Niehus, R., Hay, J., Salazar, P.D., Meakin, S., Munday, J., Bosse, N.I., Sherrat, K., Robin, N., White, L.F., Huisman, J.S., Stadler, T., Wallinga, J., Funk, S., Lipsitch, M. & Cobey, S. (2020) Practical considerations for measuring the effective reproductive number R t.

Grolemund, G. & Wickham, H. (2011) Dates and times made easy with lubridate. Journal of Statistical Software, 40, 1–25.

Hinch, R., Probert, W.J., Nurtay, A., Kendall, M., Wymant, C., Hall, M., Lythgoe, K., Cruz, A.B., Zhao, L., Stewart, A., Ferretti, L., Montero, D., Warren, J., Mather, N., Abueg, M., Wu, N., Legat, O., Bentley, K., Mead, T., Van-Vuuren, K., Feldner-Busztin, D., Ristori, T., Finkelstein, A., Bonsall, D.G., Abeler-DÃ¶rner, L. & Fraser, C. (2021) Openabm-covid19-an agent-based model for non-pharmaceutical interventions against covid-19 including contact tracing. PLoS Computational Biology, 17.

Hopkins, S.R., Fleming-Davies, A.E., Belden, L.K. & Wojdak, J.M. (2020) Systematic review of modelling assumptions and empirical evidence: Does parasite transmission increase nonlinearly with host density? Methods in Ecology and Evolution, 11, 476–486.

Hou, X., Gao, S., Li, Q., Kang, Y., Chen, N., Chen, K., Rao, J., Ellenberg, J.S. & Patz, J.A. (2021) Intracounty modeling of covid-19 infection with human mobility: Assessing spatial heterogeneity with business traffic, age, and race. Proceedings of the National Academy of Sciences, 118.

Hu, T., Wang, S., She, B., Zhang, M., Huang, X., Cui, Y., Khuri, J., Hu, Y., Fu, X., Wang, X., Wang, P., Zhu, X., Bao, S., Guan, W. & Li, Z. (2021) Human mobility data in the covid-19 pandemic: characteristics, applications, and challenges. International Journal of Digital Earth, 14, 1126–1147.

Hwang, K.K.L., Edholm, C.J., Saucedo, O., Allen, L.J.S. & Shakiba, N. (2022) A hybrid epidemic model to explore stochasticity in covid-19 dynamics. Bulletin of Mathematical Biology, 84, 91.

Keeling, M. & Rohani, P. (2008) Modeling infectious diseases in humans and animals. Princeton University Press.

Kerr, C.C., Stuart, R.M., Mistry, D., Abeysuriya, R.G., Rosenfeld, K., Hart, G.R., NÃºÃ±ez, R.C., Cohen, J.A., Selvaraj, P., Hagedorn, B., George, L., JastrzÈ©bski, M., Izzo, A.S., Fowler, G., Palmer, A., Delport, D., Scott, N., Kelly, S.L., Bennette, C.S., Wagner, B.G., Chang, S.T., Oron, A.P., Wenger, E.A., Panovska-Griffiths, J., Famulare, M. & Klein, D.J. (2021) Covasim: An agent-based model of covid-19 dynamics and interventions. PLoS Computational Biology, 17.

Kraemer, M.U., Golding, N., Bisanzio, D., Bhatt, S., Pigott, D.M., Ray, S.E., Brady, O.J., Brownstein, J.S., Faria, N.R., Cummings, D.A., Pybus, O.G., Smith, D.L., Tatem, A.J., Hay, S.I. & Reiner, R.C. (2019) Utilizing general human movement models to predict the spread of emerging infectious diseases in resource poor settings. Scientific Reports, 9.

Lachiany, M. & Stone, L. (2012) A Vaccination Model for a Multi-City System. Bulletin of Mathematical Biology, 74, 2474–2487.

Lambert, S., Ezanno, P., Garel, M. & Gilot-Fromont, E. (2018) Demographic stochasticity drives epidemiological patterns in wildlife with implications for diseases and population management. Scientific Reports, 8.

Lavine, J.S., Bjornstad, O.N. & Antia, R. (2021) Immunological characteristics govern the transition of covid-19 to endemicity. Science, 371, 741–745.

Lloyd-Smith, J.O., Schreiber, S.J., Kopp, P.E. & Getz, W.M. (2005) Superspreading and the effect of individual variation on disease emergence. Nature, 438, 355.

Metcalf, C.J., Edmunds, W.J. & Lessler, J. (2015) Six challenges in modelling for public health policy. Epidemics, 10, 93–96.

Pan, A., Liu, L., Wang, C., Guo, H., Hao, X., Wang, Q., Huang, J., He, N., Yu, H., Lin, X., Wei, S. & Wu, T. (2020) Association of public health interventions with the epidemiology of the covid-19 outbreak in wuhan, china. JAMA, 323, 1915.

Pei, S., Kandula, S. & Shaman, J. (2020) Differential effects of intervention timing on covid-19 spread in the united states.

Riley, S. (2007) Large-Scale Models of Infectious Disease. Science, 316, 1298–1301.

Rohani, P., Earn, D. & Grenfell, B.T. (1999) Opposite Patterns of Synchrony in Sympatric Disease Metapopulations. Science, 286, 968–971.

Saad-Roy, C.M., Wagner, C.E., Baker, R.E., Morris, S.E., Farrar, J., Graham, A.L., Levin, S.A., Mina, M.J., Metcalf, C.J.E. & Grenfell, B.T. (2020) Immune life history, vaccination, and the dynamics of sars-cov-2 over the next 5 years. Science, 370, 811–818.

Shea, K., Runge, M.C., Pannell, D., Probert, W.J.M., Li, S.L., Tildesley, M. & Ferrari, M. (2020) Harnessing multiple models for outbreak management. Science, 368, 577–579.

Tian, H., Liu, Y., Li, Y., Wu, C.H., Chen, B., Kraemer, M.U., Li, B., Cai, J., Xu, B., Yang, Q. et al. (2020) An investigation of transmission control measures during the first 50 days of the covid-19 epidemic in china. Science, 368, 638–642.

Wang, L. & Li, X. (2014) Spatial epidemiology of networked metapopulation: an overview. Chinese Science Bulletin, 59, 3511–3522.

Wardle, J., Bhatia, S., Kraemer, M.U.G., Nouvellet, P. & Cori, A. (2022) Gaps in mobility data and implications for modelling epidemic spread: a scoping review and simulation study. medRxiv.

Yamana, T., Pei, S. & Shaman, J. (2020) Projection of COVID-19 Cases and Deaths in the US as Individual States Re-open May 4,2020. medRxiv, p. 2020.05.04.20090670.

Yang, W., Kandula, S., Huynh, M., Greene, S.K., Wye, G.V., Li, W., Chan, H.T., McGibbon, E., Yeung, A., Olson, D., Fine, A. & Shaman, J. (2021) Estimating the infection-fatality risk of sars-cov-2 in new york city during the spring 2020 pandemic wave: a model-based analysis. The Lancet Infectious Diseases, 21, 203–212.

Zebrowski, A., Rundle, A., Pei, S., Yaman, T., Yang, W., Carr, B.G., Sims, S., Doorley, R., Schluger, N., Quinn, J.W., Shaman, J. & Branas, C.C. (2021) A spatiotemporal tool to project hospital critical care capacity and mortality from covid-19 in us counties. American journal of public health, 111, 1113–1122.

Zhang, M., Wang, S., Hu, T., Fu, X., Wang, X., Hu, Y., Halloran, B., Li, Z., Cui, Y., Liu, H., Liu, Z. & Bao, S. (2022) Human mobility and covid-19 transmission: a systematic review and future directions. Annals of GIS.

